# Mucosal Correlates of Protection after Influenza Viral Challenge of Vaccinated and Unvaccinated Healthy Volunteers

**DOI:** 10.1101/2023.09.27.23296227

**Authors:** Rachel Bean, Luca T. Giurgea, Alison Han, Lindsay Czajkowski, Adriana Cervantes-Medina, Monica Gouzoulis, Allyson Mateja, Sally Hunsberger, Susan Reed, Rani Athota, Holly Ann Baus, John C. Kash, Jaekeun Park, Jeffery K. Taubenberger, Matthew J. Memoli

**Affiliations:** LID Clinical Studies Unit, Laboratory of Infectious Diseases, National Institute of Allergy and Infectious Diseases, National Institutes of Health, Bethesda, Maryland, USA; Clinical Monitoring Research Program Directorate, Frederick National Laboratory for Cancer Research, Frederick, Maryland, USA; Biostatistics Research Branch, National Institute of Allergy and Infectious Diseases, National Institutes of Health, Bethesda, Maryland, USA; Viral Pathogenesis and Evolution Section, Laboratory of Infectious Diseases, National Institute of Allergy and Infectious Diseases, National Institutes of Health, Bethesda, Maryland, USA; Department of Veterinary Medicine, VA-MD College of Veterinary Medicine, University of Maryland, College Park, MD

## Abstract

Induction of systemic antibody titers against hemagglutinin has long been the main focus of influenza vaccination strategies, but mucosal immunity has also been shown to play a key role in protection against respiratory viruses. By vaccinating and challenging healthy volunteers, we demonstrated that inactivated influenza vaccine (IIV) modestly reduced the rate of influenza while predominantly boosting serum antibody titers against hemagglutinin (HA) and HA stalk, a consequence of the low neuraminidase (NA) content of IIV and the intramuscular route of administration. Not surprisingly, viral challenge induced nasal and serum responses against both HA and NA. Correlations between mucosal IgA and serum IgG against specific antigens were low, whether before or after challenge, suggesting a compartmentalization of immune responses. Even so, volunteers who developed viral shedding for multiple days had lower baseline titers across both systemic and mucosal compartments as compared to those with no shedding or a single day of shedding. Regression analysis showed that pre-challenge HA inhibition titers were the most consistent correlate of protection across clinical outcomes combining shedding and symptoms, with NA inhibition titers and HA IgG levels only predicting the duration of shedding. Despite the inclusion of data from multiple binding and functional antibody assays against HA and NA performed on both serum and nasal samples, multivariate models were unable to account for the variability in outcomes, emphasizing our imperfect understanding of immune correlates in influenza and the importance of refining models with assessments of innate and cellular immune responses.

**Importance:** The devastating potential of influenza has been well known for over 100 years. Despite the development of vaccines since the middle of the twentieth century, influenza continues to be responsible for substantial global morbidity and mortality. To develop next-generation vaccines with enhanced effectiveness, we must synthesize our understanding of the complex immune mechanisms culminating in protection. Our study outlines the differences in immune responses to influenza vaccine and influenza infection, identifying potential gaps in vaccine-induced immunity, particularly at the level of the nasal mucosa. Furthermore, this research underscores the need to refine our imperfect models while recognizing potential pitfalls in past and future attempts to identify and measure correlates of protection.

## Introduction

Induction of robust, durable immune protection against RNA respiratory viruses such as influenza poses a significant scientific and public health challenge (1). Vaccination is a fundamental component of public health responses, yet the efficacy of current vaccine strategies is limited by viral immune evasion adaptations and waning host immunity. Contemporary vaccines are unable to effectively prevent spread of the virus and often offer inadequate protection to individuals at the highest risk of complications and death. The development of new vaccine strategies offering more robust, broad, and durable protection is urgently needed, but requires an improvement in our understanding of immunoprotection.

Existing influenza vaccines induce a systemic immune response to the major surface protein, hemagglutinin (HA), and are standardized by stimulation of serum anti-HA antibodies as the primary correlate of protection. The FDA and EMA CHMP both define “protective titers” as a serum HAI titer of ≥ 40 (2). Despite this acceptance of HA-specific antibodies as a surrogate for immunity, there is a body of research highlighting the importance of the immune response to other antigenic targets, such as neuraminidase (NA) (3–7). The role of mucosal immunity in influenza A virus infection has long been studied (8), and recent research on mucosal immunity against both influenza and SARS-COV-2 has illustrated the significance of nasal IgA in protection against respiratory viruses (9–12). Stimulation of robust mucosal immunity has shown promise as a mechanism to reduce viral transmission, an outcome that is largely unaffected by systemically administered vaccines (13–15). Further potential benefits of strategies targeting mucosal immunity include more potent viral neutralization via polymeric secretory IgA (16, 17) and improved durability of responses (18). Intranasally delivered live-attenuated influenza vaccine has demonstrated a capacity to provide superior efficacy compared to intramuscular vaccine, though inconsistently (19, 20). Hence, the ideal amount, kinetics, and antigenic specificity of mucosal antibodies needed to confer protection from influenza remain unknown.

In the setting of this public health and scientific impetus, this trial was designed to investigate mucosal and systemic immunity after influenza vaccination and infection in a human challenge study. This initial report summarizes the immune responses to vaccination and infection, describes the relationship between mucosal and systemic immunity, explores the correlation of immune responses to clinical outcomes, and raises questions that must be addressed as we pursue more protective vaccines against respiratory viruses.

## Results

### Study Population

The study enrolled a total of 80 participants, 40 who were vaccinated intramuscularly (IM) with standard quadrivalent influenza vaccine (IIV) and 40 who remained unvaccinated. Three participants in the vaccinated cohort withdrew prior to viral challenge, 2 due to voluntary withdrawal and 1 due to safety lab abnormalities. Three in the unvaccinated cohort withdrew prior to viral challenge; 2 voluntarily withdrew, and 1 had a positive nasal wash for another respiratory virus prior to challenge. Therefore, 74 participants (37 per cohort) were challenged with influenza and included in the final analysis. The average age of participants was 34 years. Similar distributions of age, race, gender, and ethnicity were enrolled in both the vaccinated and unvaccinated cohorts (Table 1).

**Table 1.**
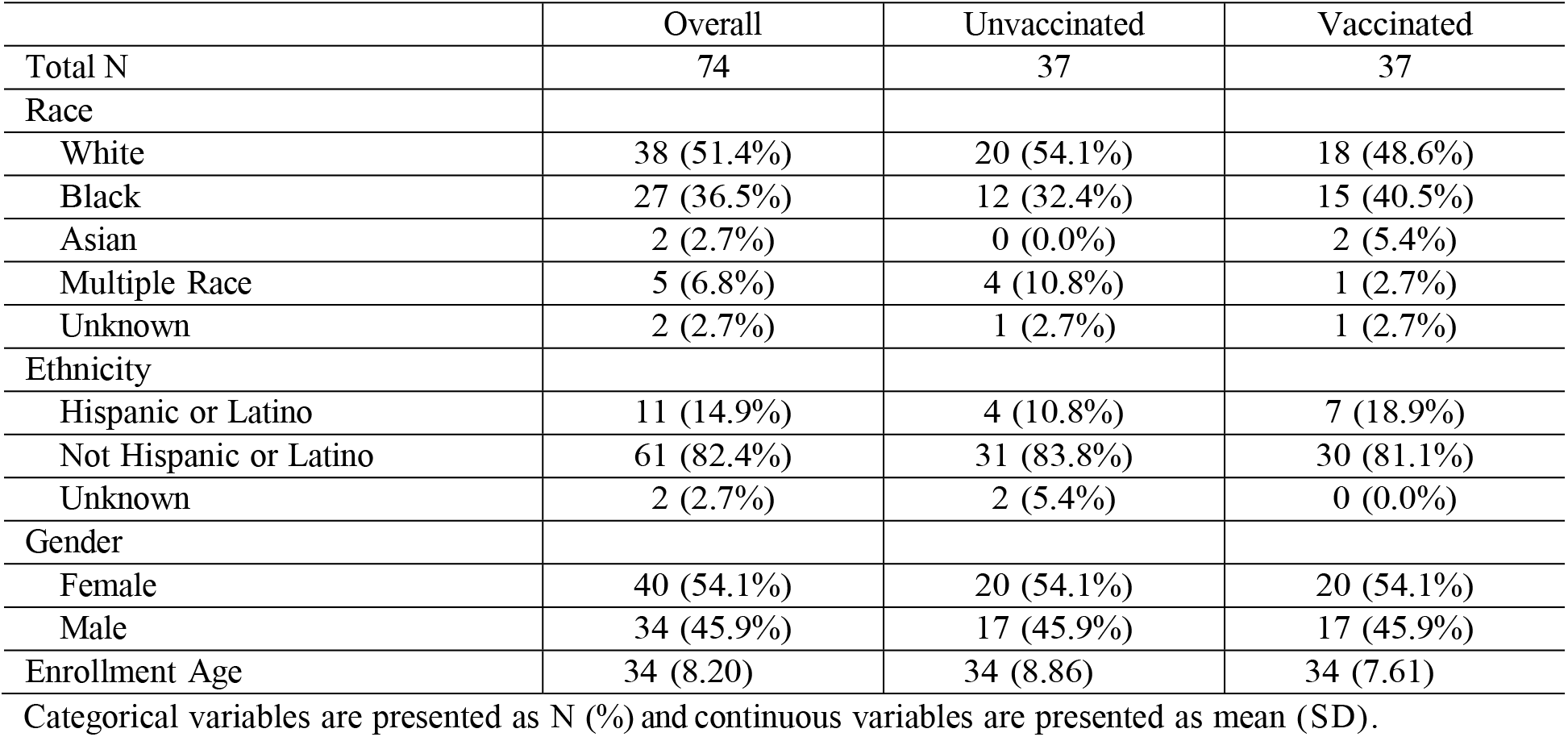
Demographics of Participants.

|  | Overall | Unvaccinated | Vaccinated |
| --- | --- | --- | --- |
| Total N | 74 | 37 | 37 |
| Race |  |  |  |
| White | 38 (51.4%) | 20 (54.1%) | 18 (48.6%) |
| Black | 27 (36.5%) | 12 (32.4%) | 15 (40.5%) |
| Asian | 2 (2.7%) | 0 (0.0%) | 2 (5.4%) |
| Multiple Race | 5 (6.8%) | 4 (10.8%) | 1 (2.7%) |
| Unknown | 2 (2.7%) | 1 (2.7%) | 1 (2.7%) |
| Ethnicity |  |  |  |
| Hispanic or Latino | 11 (14.9%) | 4 (10.8%) | 7 (18.9%) |
| Not Hispanic or Latino | 61 (82.4%) | 31 (83.8%) | 30 (81.1%) |
| Unknown | 2 (2.7%) | 2 (5.4%) | 0 (0.0%) |
| Gender |  |  |  |
| Female | 40 (54.1%) | 20 (54.1%) | 20 (54.1%) |
| Male | 34 (45.9%) | 17 (45.9%) | 17 (45.9%) |
| Enrollment Age | 34 (8.20) | 34 (8.86) | 34 (7.61) |

Categorical variables are presented as N (%) and continuous variables are presented as mean (SD).

### Clinical Outcomes

The binary clinical outcomes of the viral challenge are summarized in Table 2. Compared to the unvaccinated cohort, a significantly lower proportion of the vaccinated cohort developed Flu (defined as at least one symptom, plus either at least one day of viral shedding or a ≥4-fold increase in either HAI or NAI titers), mild to moderate influenza disease (MMID, defined as at least one symptom plus at least one day of viral shedding), and symptoms. While shedding was less common in vaccinated participants, this difference was not statistically significant. In the vaccinated vs. unvaccinated cohort, trends towards shorter duration of shedding, shorter duration of symptoms, and lower number of symptoms were observed (p=0.0944, p=0.0819, and p=0.0634, respectively) (Figure 1, Table S2). InFLUenza Patient-Reported Outcome (FLU-PRO©) scores, a validated measure of symptom severity, were lower in the vaccinated cohort (p=0.0434) (Figure 1, Table S2) (21).

**Fig 1.**
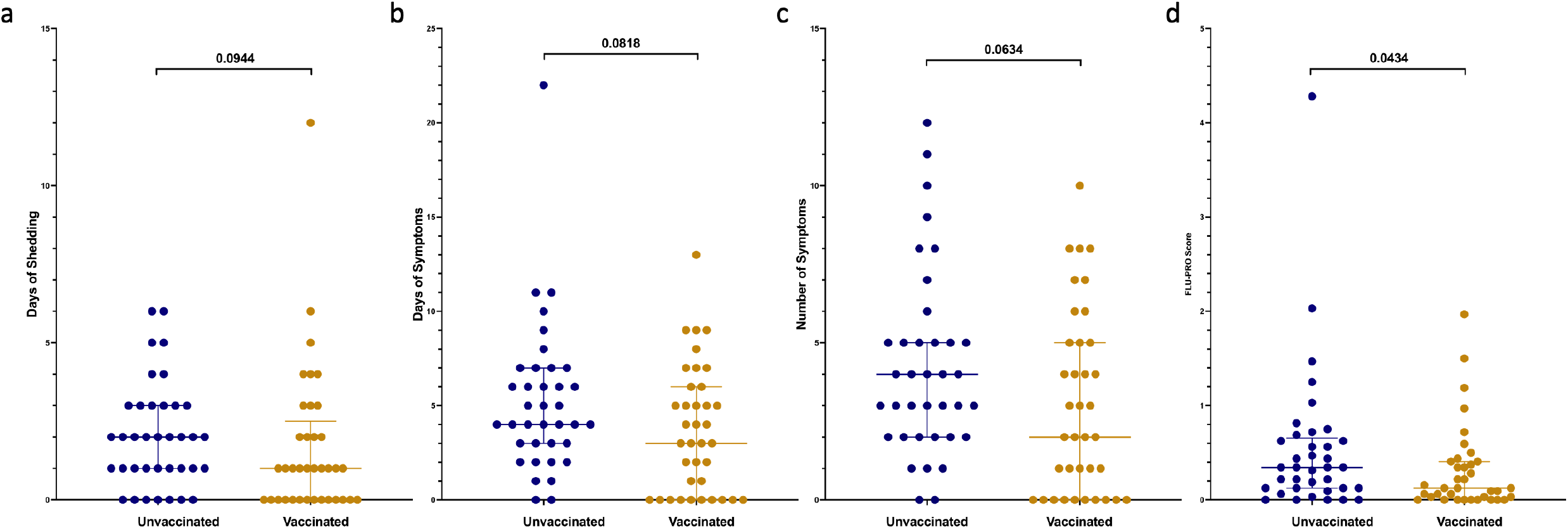
Clinical outcomes after challenge demonstrating duration (days) of shedding (A), duration (days) of symptoms (B), total number of symptoms (C), and FLU-PRO (D) scores, which standardize symptom severity based on number of symptoms and duration with higher numbers signifying more severe illness. Individual values represented by dots for unvaccinated participants in blue and vaccinated participants in yellow. Horizontal lines represent medians and first and third quartiles. P-values from Wilcoxon rank sum tests comparing unvaccinated and vaccinated participants shown, with p<0.05 considered statistically significant.

**Table 2.** Incidence of Binary Clinical Outcomes after H1N1 Challenge.

|  | Unvaccinated N (%) | Vaccinated N (%) | P-Value<sup>a</sup> |
| --- | --- | --- | --- |
| Total N | 37 | 37 |  |
| MMID<sup>b</sup> | 30 (81%) | 18 (49%) | 0.0068 |
| Flu<sup>c</sup> | 34 (92%) | 22 (59%) | 0.0023 |
| Symptoms<sup>d</sup> | 35 (95%) | 27 (73%) | 0.0242 |
| Shedding<sup>e</sup> | 30 (81%) | 23 (62%) | 0.1208 |

<sup>a</sup>P-values from Fisher's Exact Tests with no adjustment for multiple comparisons
<sup>b</sup>MMID (mild to moderate influenza disease) defined as at least 1 influenza symptom plus at least 1 day of viral shedding
<sup>c</sup>Flu defined as at least 1 influenza symptom, plus either at least 1 day of viral shedding or a $\geq 4$ fold increase in either hemagglutinin inhibition titers or neuraminidase inhibition titers
<sup>d</sup>Symptoms defined as presence of at least 1 influenza symptom
<sup>e</sup>Shedding defined as presence of shedding on at least 1 day

### Serum and Nasal Humoral Immune Responses

Antibody titers from serum and nasal mucosa were assessed pre– and post-challenge in both cohorts, and in the vaccinated cohort, pre– and post-vaccination. Twenty-eight days after vaccination, significant increases in serum anti-HA IgG and anti-HA stalk IgG ELISA titers (Figure 4) were observed (p<0.0001 for both), as well as a modest increase in serum HAI titers that was not statistically significant (Figure 2). However, no increase was observed in serum antibodies against NA, or in mucosal IgA titers against HA, HA stalk or NA (Figure 2, 3, 4).

**Fig 2.**
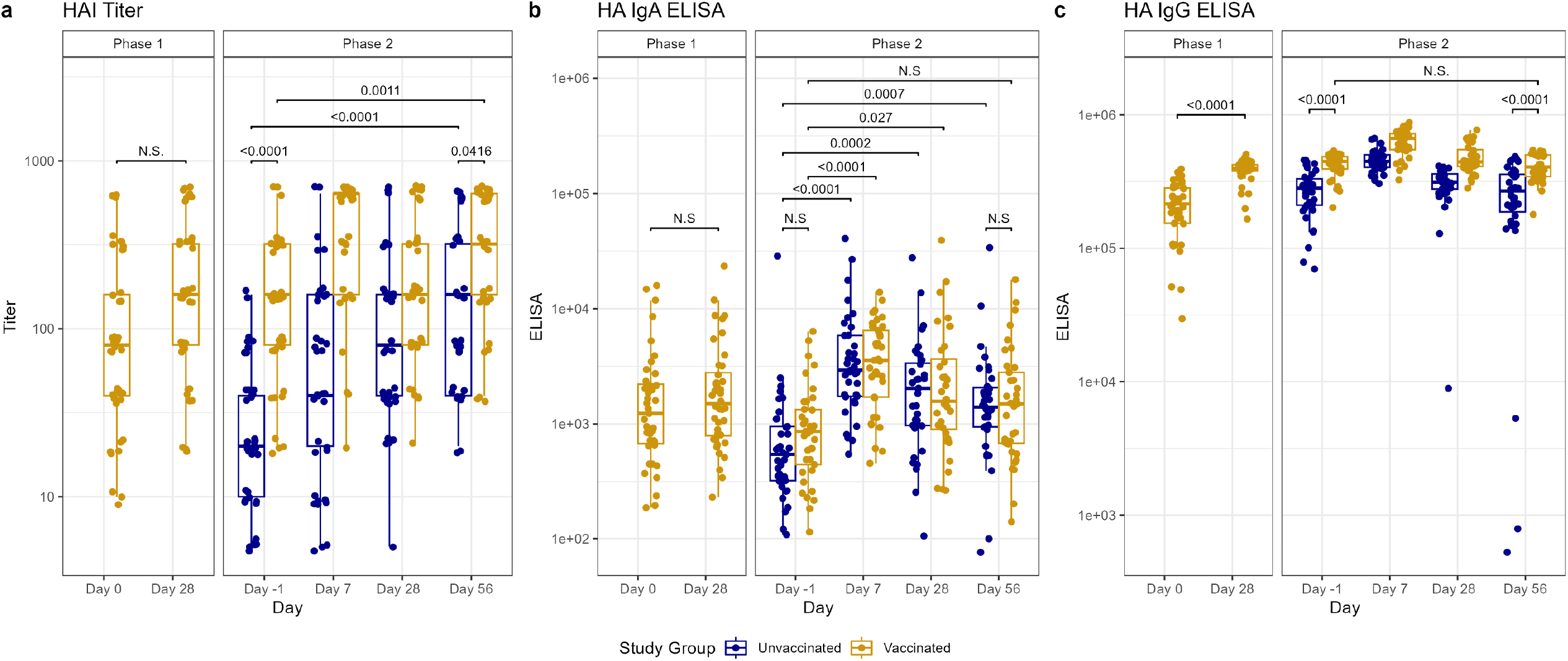
Antibody titers after vaccination (Phase 1) and H1N1 viral challenge (Phase 2) as tested by hemagglutinin (HA) inhibition (22) assays (A), ELISA for nasal anti-HA IgA (B), and ELISA for serum anti-HA IgG (C). In phase 1, day 0 represents baseline titers on the day of vaccination. Subsequently in phase 2, day –1 represents baseline titers prior to viral challenge on day 0. Individual values represented by dots for unvaccinated participants in blue and vaccinated participants in yellow. Horizontal lines represent medians and first and third quartiles. Vertical lines represent minimum and maximum non-outliers. P-values from Wilcoxon rank sum tests, adjusted for multiple comparisons by Holm’s method, comparing unvaccinated and vaccinated participants shown, with p<0.05 considered statistically significant. Important non-significant (N.S.) comparisons also shown.

**Fig 3.**
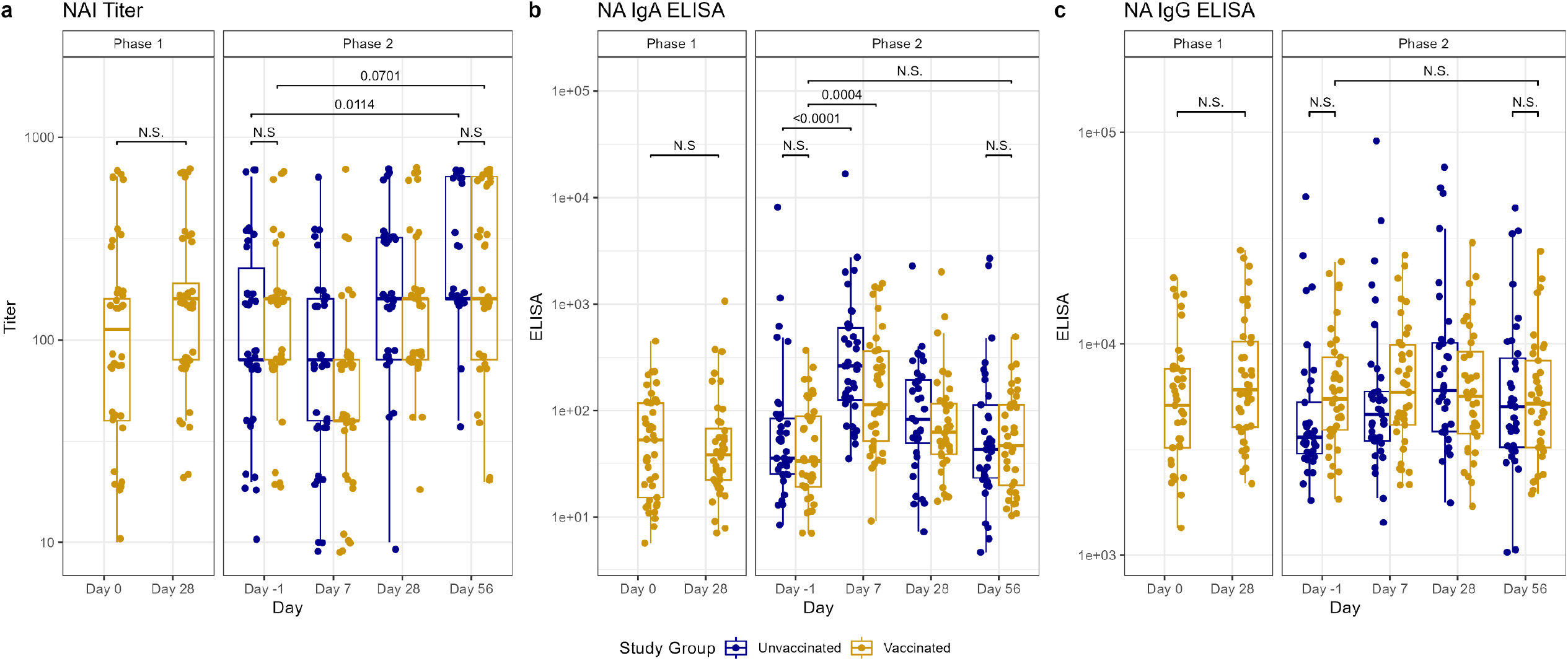
Antibody titers after vaccination (Phase 1) and H1N1 viral challenge (Phase 2) as tested by neuraminidase (NA) inhibition (NAI) assays (A), ELISA for nasal anti-NA IgA (B), and ELISA for serum anti-NA IgG (C). In phase 1, day 0 represents baseline titers on the day of vaccination. Subsequently in phase 2, day –1 represents baseline titers prior to viral challenge on day 0. Individual values represented by dots for unvaccinated participants in blue and vaccinated participants in yellow. Horizontal lines represent medians and first and third quartiles. Vertical lines represent minimum and maximum non-outliers. P-values from Wilcoxon rank sum tests, adjusted for multiple comparisons by Holm’s method, comparing unvaccinated and vaccinated participants shown, with p<0.05 considered statistically significant. Important non-significant (N.S.) comparisons also shown.

**Fig 4.**
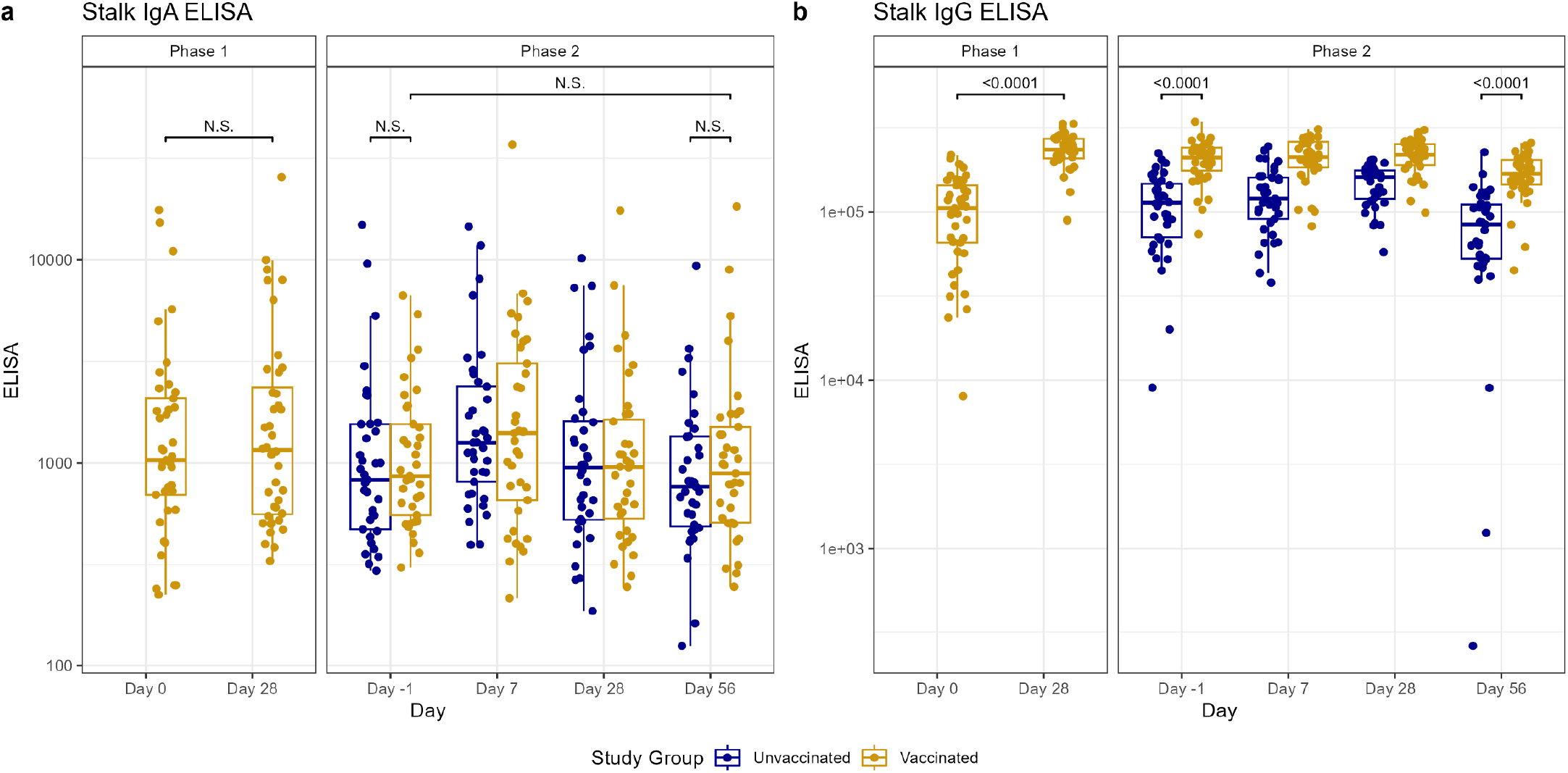
Antibody titers after vaccination (Phase 1) and H1N1 viral challenge (Phase 2) as tested by ELISA for nasal anti-HA stalk IgA (A) and ELISA for serum anti-HA stalk IgG (B). In phase 1, day 0 represents baseline titers on the day of vaccination. Subsequently in phase 2, day –1 represents baseline titers prior to viral challenge on day 0. Individual values represented by dots for unvaccinated participants in blue and vaccinated participants in yellow. Horizontal lines represent medians and first and third quartiles. Vertical lines represent minimum and maximum non-outliers. P-values from Wilcoxon rank sum tests, adjusted for multiple comparisons by Holm’s method, comparing unvaccinated and vaccinated participants shown, with p<0.05 considered statistically significant. Important non-significant (N.S.) comparisons also shown.

The vaccinated cohort demonstrated rises in HA inhibition (22) titers between day –1 pre-challenge and day 56 post-challenge (p=0.0011), despite starting with a significantly higher HAI titer than the unvaccinated cohort on day –1 (p<0.0001) (Figure 2). HAI titers also increased by day 56 post-challenge, compared to day –1 pre-challenge, in the unvaccinated cohort (p<0.0001) (Figure 2), with more individuals developing a ≥4-fold rise in titer than in the vaccinated cohort (29 vs. 15, p=0.0001) (Table S2). However, we observed an overall higher HAI titer in the vaccinated cohort on day 56 than in the unvaccinated (p=0.0416) (Figure 2). There was also an increase in NAI titers on day 56 post-challenge compared to day –1 pre-challenge in both the unvaccinated cohort (p=0.0114) and the vaccinated cohort (p=0.0701), though the latter did not reach statistical significance. No difference in mean NA inhibition (NAI) titer was noted between the cohorts on day 56 (Figure 3).

Prior to challenge, vaccinated participants had higher anti-HA IgG and anti-HA stalk serum IgG (p<0.0001 for both) compared to unvaccinated participants. There were no between-group differences in the other day –1 serum or any of the mucosal antibody titers from the vaccinated and unvaccinated cohorts (Figure 2, Figure 3, Figure 4). In the vaccinated cohort, there was an increase in mucosal anti-HA IgA and anti-NA IgA on day 7 (p<0.0001, 0.0004, respectively) and mucosal anti-HA IgA on day 28 (p=0.027) after challenge. In unvaccinated individuals, we observed similar rises in mucosal anti-HA IgA and anti-NA IgA titers by day 7 (p<0.0001, <0.0001, respectively), and we observed the rise in anti-HA IgA to be sustained longer, at both day 28 (p=0.0002) and day 56 (p=0.0007) (Figure 2, Figure 3). Overall, on day 56 post-challenge, the vaccinated cohort demonstrated higher levels of anti-HA IgG and anti-Stalk IgG in the serum (p<0.0001, <0.0001, respectively) compared to the unvaccinated. No other serum or mucosal ELISA titers were significantly different between study cohorts on day 56 post-challenge. When comparing the change in titer levels from day –1 pre-challenge to other study timepoints between vaccinated and unvaccinated cohorts, only the change from day –1 pre-challenge to day 56 post challenge in serum anti-NA IgG was significantly different (p=0.0032), with unvaccinated subjects having a larger increase compared to vaccinated (Figure 2, Figure 3, Figure 4, Table S3).

### Correlations between Serum and Nasal Humoral Immune Responses

For all participants regardless of vaccination cohort, the correlations between antibody titers were generated at multiple timepoints (pre-challenge, 7 days post-challenge, and 56 days post-challenge), and the correlations were generally similar across timepoints except where noted. Serum HAI titers were highly correlated to serum HA IgG and, to a lesser degree, HA stalk IgG, but not NA IgG (Figure 5). Serum NAI titers were moderately correlated to serum NA IgG titers (significantly at day –1 and day 7 but not day 56), but not to serum HA IgG or HA stalk IgG. Serum HA IgG correlated well with serum HA stalk IgG pre-challenge, and serum NA IgG generally did not correlate well with HA IgG (except at day 56, rho 0.411) or HA stalk IgG. Nasal HA IgA, HA stalk IgA, and NA IgA correlated moderately well with each other. Notably, serum IgG titers against each antigen had no correlation with nasal IgA titers against the same antigen (e.g., serum HA IgG and nasal HA IgA). None of the antigen-specific antibody titers demonstrated correlation with total serum IgG or IgA levels. Serum antibody titers against specific antigens had high correlations across timepoints (e.g., HA IgG at day –1 compared to HA IgG at day 7) and were highly statistically significant (Table S4). In contrast, mucosal antibody responses across timepoints correlated less strongly (Table S4).

**Fig 5.**
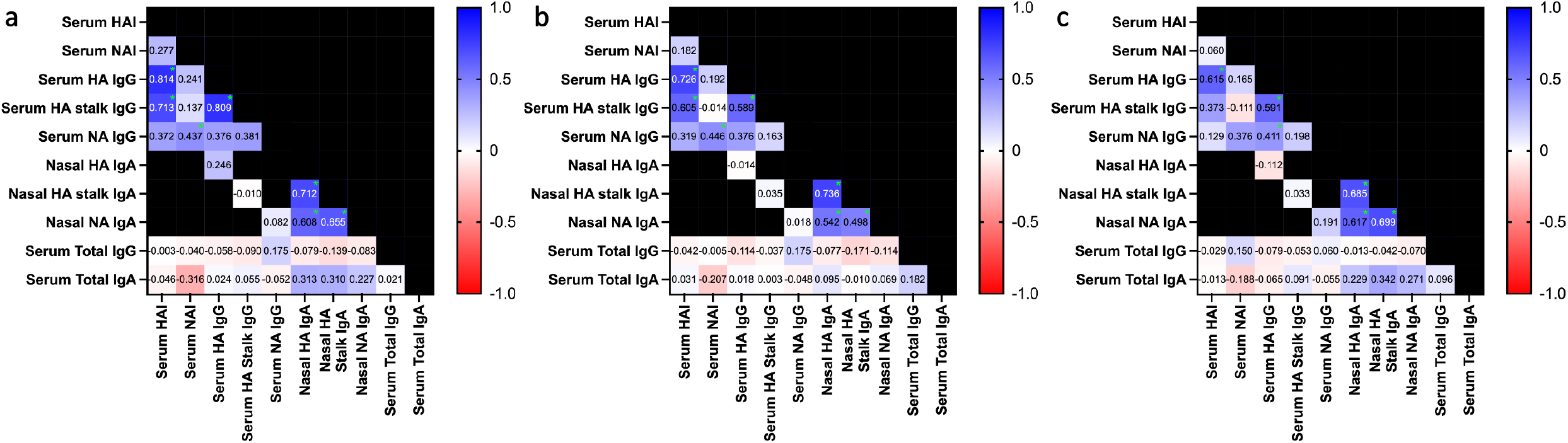
Heatmaps of spearman rank correlation coefficients between serum and nasal antibody titers. Numbers within squares and square colors represent rho values, with blue indicating positive and red representing negative correlations. Green asterisks indicate statistically significant comparisons after Bonferroni adjustment. Black squares represent comparisons left untested for scientific or statistical reasons.

### Comparisons of Clinical Outcomes and Humoral Responses by Shedding Status

Grouping participants by duration of shedding (non-shedders, 1-day shedders, long-shedders) demonstrated significant differences in multiple antibody titers across groups, with the highest titers seen in non-shedders and 1-day shedders and the lowest in long-shedders, for serum HAI, serum NAI, serum HA IgG, and nasal HA IgA. Antibody titers were not significantly different between non-shedders and 1-day shedders (Table 3, Table S5). The proportion of symptoms and number of symptoms were different across groups, but pairwise differences were not statistically significant when adjusted for multiple comparisons (Table 4).

**Table 3.**
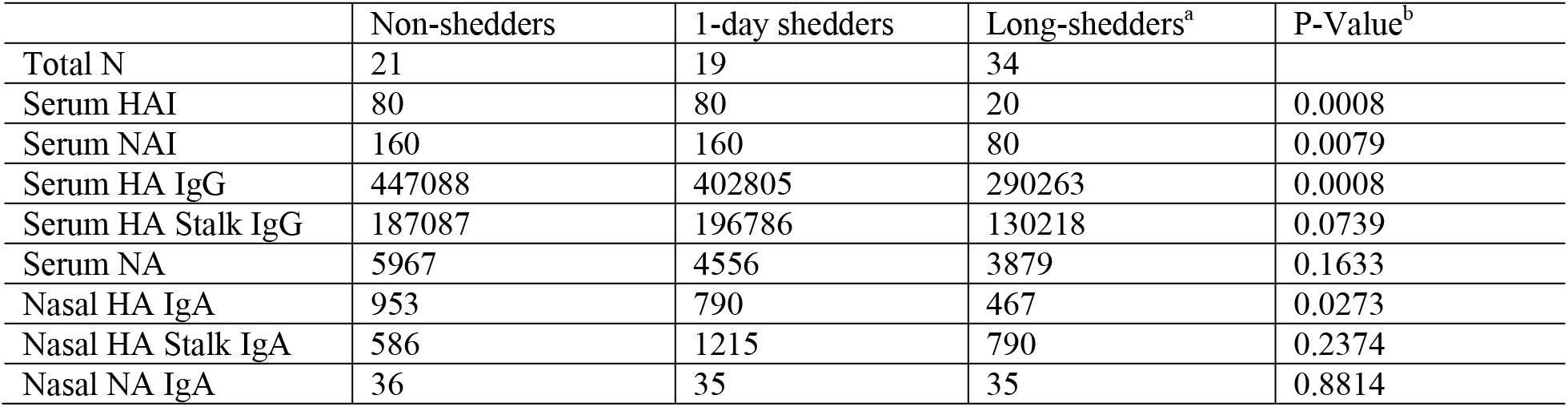
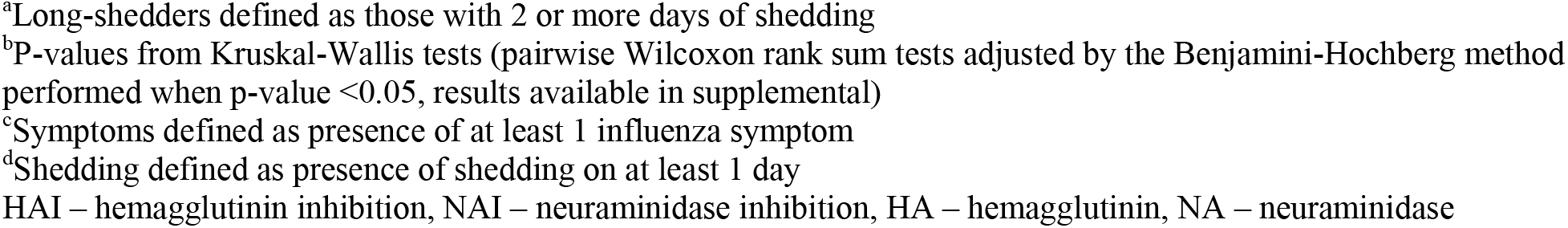
Median Antibody Titers before H1N1 Challenge at Day –1 Grouped by Participant Shedding Status.

|  | Non-shedders | 1-day shedders | Long-shedders<sup>a</sup> | P-Value<sup>b</sup> |
| --- | --- | --- | --- | --- |
| Total N | 21 | 19 | 34 |  |
| Serum HAI | 80 | 80 | 20 | 0.0008 |
| Serum NAI | 160 | 160 | 80 | 0.0079 |
| Serum HA IgG | 447088 | 402805 | 290263 | 0.0008 |
| Serum HA Stalk IgG | 187087 | 196786 | 130218 | 0.0739 |
| Serum NA | 5967 | 4556 | 3879 | 0.1633 |
| Nasal HA IgA | 953 | 790 | 467 | 0.0273 |
| Nasal HA Stalk IgA | 586 | 1215 | 790 | 0.2374 |
| Nasal NA IgA | 36 | 35 | 35 | 0.8814 |

<sup>a</sup>Long-shedders defined as those with 2 or more days of shedding <sup>b</sup>P-values from Kruskal-Wallis tests (pairwise Wilcoxon rank sum tests adjusted by the Benjamini-Hochberg method performed when p-value <0.05, results available in supplemental) <sup>c</sup>Symptoms defined as presence of at least 1 influenza symptom <sup>d</sup>Shedding defined as presence of shedding on at least 1 day HAI – hemagglutinin inhibition, NAI – neuraminidase inhibition, HA – hemagglutinin, NA – neuraminidase

**Table 4.** Clinical Outcomes after H1N1 Challenge Grouped by Participant Shedding Status.

|  | Non-shedders | 1-day shedders | Long-shedders<sup>a</sup> | P-Value<sup>b</sup> |
| --- | --- | --- | --- | --- |
| Total N | 21 | 19 | 34 |  |
| Shedding<sup>c,d</sup> | 0 | 19 (100%) | 34 (100%) |  |
| Symptoms<sup>d</sup> | 14 (67%) | 18 (95%) | 30 (88%) | 0.0350 |
| MMID<sup>c,e</sup> | 0 | 18 (95%) | 30 (88%) |  |
| Flu<sup>c,f</sup> | 8 (38%) | 18 (95%) | 30 (88%) |  |
| Median (IQR) Days of Shedding<sup>c</sup> | 0 (0, 0) | 1 (1, 1) | 3 (2, 4) |  |
| Median (IQR) Days of Symptoms | 3 (0, 5) | 5 (3, 6.5) | 4 (2, 6.75) | 0.1363 |
| Median (IQR) Number of symptoms | 2 (0, 4) | 4 (2.5, 6.5) | 3 (2, 5) | 0.0268 |

<sup>a</sup>Long-shedders defined as those with 2 or more days of shedding <sup>b</sup>P-values from Kruskal-Wallis tests (pairwise Wilcoxon rank sum tests adjusted by the Benjamini-Hochberg method performed when p-value <0.05, results available in supplemental) <sup>c</sup>Statistical analysis not performed on shedding outcomes between groups, which are defined by shedding status, and outcomes that are dependent on shedding status (Flu, MMID) <sup>d</sup>Symptoms defined as presence of at least 1 influenza symptom <sup>e</sup>Shedding defined as presence of shedding on at least 1 day <sup>f</sup>MMID (mild to moderate influenza disease) defined as at least 1 influenza symptom plus at least 1 day of viral shedding <sup>f</sup>Flu defined as at least 1 influenza symptom, plus either at least 1 day of viral shedding or a $\geq 4$ fold increase in either hemagglutinin inhibition titers or neuraminidase inhibition titers IQR – interquartile range

### Correlates of Protection Assessment through Regression Modelling

In all participants combined, pre-challenge serum HAI titers were observed to have a significant, independent, negative association with Flu, MMID, shedding, and days of shedding (Table S6, Table S7). Serum NAI titers and HA IgG at day –1 were also independently negatively associated with days of shedding. When vaccination cohorts were evaluated separately, only HAI titer and serum anti-HA IgG at day –1 were significantly negatively associated with days of shedding in vaccinated participants. In the unvaccinated cohort, none of the studied immune measures demonstrated an ability to predict outcome measures (Table S8, Table S9).

Multivariate models were identified using the Akaike information criterion (AIC), but all of them had R^2^ values of ≤ 0.5, with the majority < 0.1 (Table S10). The best models with the largest R^2^ in each cohort related to shedding and days of shedding; of note, the models for the vaccinated cohort were more complex than those for the unvaccinated but had higher R^2^ values, indicating slightly better performance as predictive models.

## Discussion

The ability of IIV to prevent influenza in this study was modest, resulting in only a 33% difference in proportions of Flu incidence between groups. This estimate fits well with real-world measures of effectiveness, validating the challenge model as a vaccine assessment tool (23, 24). In our study, vaccination did not confer a significant reduction in incidence of shedding, suggesting that the ability of IIV to prevent transmission may be very limited. This shortcoming may exist because the vaccine, given via the IM route, provoked no measurable induction of mucosal humoral immunity, as has been observed previously with IM influenza vaccines (25–33). Additionally, our observation that IM vaccination failed to increase antibody titers against NA is unsurprising, considering that the NA content of IIV is low and not standardized (34).

Immunity induced by IM vaccination contrasted profoundly with that induced by influenza infection, which provoked significant mucosal antibody titers against HA and NA by day 7. Mucosal responses appeared short-lived, peaking 7 days after challenge, in contrast to serum HAI and NAI titers, which peaked at day 56 (despite stable or decreasing levels of serum HA IgG and NA IgG after day 7). The discordance between the kinetics of functional and binding antibody titers in serum may reflect the effect of affinity maturation. Interestingly, participants in both cohorts experienced increases in serum titers after challenge, suggesting exposure to influenza may be important in boosting vaccine-induced systemic immunity. Unvaccinated individuals narrowed the difference in HAI titers by day 56, but never fully closed the gap with vaccinated individuals. The differential timing of the peak in mucosal IgA (early in the course of infection) versus serum HAI and serum NAI titers (in the recovery phase) may reflect differing immunologic roles of these antibodies, with IgA assisting in viral clearance and serum HAI and NAI contributing to long-term protection against future infections. In contrast, serum and mucosal antibody titers against HA stalk were not affected by challenge but did increase significantly after vaccination. While the general lack of HA stalk-directed humoral response has been primarily blamed on the immunodominance of the head domain (35), the discordance in responses to virus and IIV seen in this study supports the hypothesis that steric hinderance in intact virions contributes to diminished stalk responses (36). Furthermore, IIV demonstrated some potential as an HA-stalk vaccine, considering its ability to boost antibodies against stalk. Strategies to develop HA-stalk specific vaccines should include direct comparisons to currently approved IIV, to ensure elicited responses are superior.

The relationships between antibody titers measured in this study provide additional understanding of immune responses against influenza and may help guide next-generation vaccine design. Interestingly, there was no correlation between mucosal IgA titers and serum IgG titers against specific antigens. Similar findings have been recently reported in research on COVID-19 and may reflect differences between populations of IgA-producing plasma cells (residing locally in the mucosal lamina propria) and IgG-producing plasma cells (residing at distant sites such as bone marrow) (10, 15, 37, 38). Systemic antibody titers may, therefore, not be an appropriate surrogate for mucosal antibodies. The consistent lack of correlation across post-challenge timepoints further strengthens these conclusions. These findings also suggest that a vaccination strategy targeting the respiratory mucosa could be used to complement systemically administered vaccine, thereby boosting titers in both compartments.

Strong correlations were observed among systemic titers across multiple timepoints within individuals, meaning that participants with lower titers, despite experiencing a boost with challenge, continued to have relatively lower titers after challenge. Mucosal antibody levels lack this within-subject consistency over time. Further research is necessary to determine if titer differences between individuals are due to host factors, or if they reflect differences in immunologic history, such as the number, timing, and intensity of influenza exposures.

*Post-hoc* analysis of participants grouped by duration of shedding (0, 1, or >1 days) revealed significant differences in pre-challenge antibody titers, thereby providing a glimpse into potential correlates of protection. As expected, non-shedders had the highest antibody titers and long-shedders had the lowest titers, with differences noted between serum HAI, NAI, HA IgG, and nasal HA IgA, suggesting these are important markers of protection. Notably, 1-day shedders had baseline antibody titers more like non-shedders, rather than long-term shedders.

More sophisticated, direct analysis performed through regression modelling identified a few significant associations, with HAI titers at day –1 predicting outcomes related to shedding, duration of shedding, MMID, and Flu. Notably, we did not observe an association between mucosal antibody titers and clinical outcomes, in contrast to findings from a previous influenza challenge study (12). In addition, NAI titers were not as crucially predictive of clinical outcomes as was observed in aggregate data of four previous challenge studies conducted by our group (4). However, in almost all of these previous studies, participants were required to have low baseline serum HAI titers pre-challenge, which likely decreased the impact of HAI titers and increased the relative contribution of other correlates of protection, like serum NAI and mucosal IgA. In contrast, the administration of IM vaccine in this study boosted the contribution of HAI titers to protection and consequently decreased the signal from other immune correlates (39).

In a similar study performed in 1986 by Clements, *et al*., participants were given intranasal (IN) live attenuated influenza vaccination (LAIV) or IM IIV and then challenged with influenza A virus. In the IN LAIV cohort, pre-challenge nasal IgA against HA (as well as serum NAI) correlated with reductions in viral shedding and clinical illness, while in the IIV cohort, only serum-related markers (pre-challenge serum HAI, serum NAI and nasal HA IgG [which results from serum HA IgG spillover]) correlated with these outcomes (40). In our study, IM vaccination predominantly increased serum HAI titers, which emerged as the main correlate of protection. When performing regression analyses separately in the vaccinated and unvaccinated cohorts, HAI titers remained significantly predictive only for duration of shedding in the vaccinated cohort and were not predictive of any outcomes in the unvaccinated cohort. Upon further analysis with multivariate models evaluated using AIC, we found that the models were extremely limited with no R^2^ value better than 0.5 and most < 0.1. Therefore, even after developing models with data representing multi-antigen humoral responses in both serum and mucosa, we were unable to accurately predict most of the variation in outcomes due to influenza infection.

There are several limitations to this study. In particular, the small sample size limited our power to detect small differences. The study was not blinded, which may have introduced bias into the reporting of symptoms. In addition, enrollment was not randomized, instead following a pragmatic sequential enrollment approach; selection bias may have contributed to differences in baseline titers between the vaccinated and unvaccinated cohorts. It is important to note that most participants enrolled into the unvaccinated cohort were found to have low (<40) baseline HAI titers. In contrast, the vaccinated cohort had much higher baseline HAI titers, even before vaccination, suggesting potential differences in exposure to influenza or vaccination history. While the study showed short-lived mucosal responses, functional antibody assays were not performed on mucosal samples for comparison to functional antibody assays on serum; thus, the evaluation of mucosal antibodies may be considered less robust. Measurements of cellular immunity and cytokine levels were not available at this time, but these evaluations will be conducted in the future and may further refine our models. Furthermore, additional approaches such as repertoire sequencing, transcriptomics, and machine learning strategies may advance our understanding of the correlates of protection against influenza and guide next-generation vaccine development.

## Conclusion

The initial results from this study demonstrate that the correlates of protection to influenza are complex and reach beyond systemic or even mucosal antibody titers. These measures, though useful, can only predict clinical outcomes to a limited degree and do not account for the multiple immunological mechanisms underlying protection. For decades our influenza vaccination strategy has targeted the systemic humoral response, namely HAI titers; however, it is clear from this and other studies that we must expand our investigations to encompass the interaction between mucosal and systemic immunity and explore how vaccination in one immune compartment could beneficially or detrimentally effect the other. Further analysis of samples from studies like this will be required to elucidate how the immune system orchestrates cellular, innate, and humoral responses, within both the systemic and mucosal compartments, to combat infection and how we can translate this knowledge into better protection against disease.

## Materials and Methods

### Study Design

The challenge study was performed at the NIH Clinical Center between April and October of 2019 after participants signed consent. The primary objective was to identify mucosal correlates of protection against influenza in vaccinated and unvaccinated healthy volunteers. Protection was evaluated using a primary endpoint of Flu defined as at least one symptom of influenza plus either 1) a positive clinical test for influenza within 10 days of inoculation or 2) a fourfold or greater increase in anti-HA or anti-NA antibody titer at 56 days after administration of challenge virus. Secondary protection endpoints included incidence of shedding, incidence of symptoms, duration of shedding, duration and number of symptoms, FLU-PRO score, and incidence of MMID, defined as viral shedding detected by clinical molecular testing plus a minimum of one symptom of influenza after intranasal challenge, as previously described (41). This study (clinicaltrials.gov identifier NCT01971255) was approved by the NIAID Institutional Review Board and was conducted in accordance with the provisions of the Declaration of Helsinki and Good Clinical Practice guidelines.

The first 40 participants were enrolled in the vaccinated cohort and were vaccinated IM with IIV (Flucelvax®), referred to as phase 1 of the study. Mucosal cell samples were collected from the middle turbinate using a nasal speculum and pathology brush. Nasal secretion samples were collected using a synthetic absorbent material (SAM) strip. Mucosal samples, whole blood, PBMCs, and serum were collected pre-vaccination on the day of vaccination as well as on day 3 and day 28 post-vaccination. All vaccinations in this cohort were administered 32-155 days prior to challenge (median 64 days). Another 40 participants were identified and enrolled in the unvaccinated cohort, and 74 participants from both cohorts were brought into the NIH Clinical Center as mixed cohorts for challenge with influenza A virus.

All participants were challenged with 10^7^ TCID_50_ of the Influenza A/Bethesda/MM2/H1N1 challenge virus and were assessed daily for a minimum of 9 days in the isolation unit as previously described, referred to as phase 2 of the study (41). Mucosal and systemic samples were collected pre-challenge (the day prior to inoculation) and on days 1, 3, 5, and 7 post-challenge. Samples were also collected on day 28 and day 56 post-challenge during outpatient follow-up.

### Vaccine

The licensed 2018-2019 northern hemisphere version of Flucelvax® Quadrivalent, produced by Seqirus (Holly Springs, North Carolina), was administered to the vaccinated cohort. It contained 15 micrograms of each of 4 virus HA subunits including H3N2, H1N1, and two Influenza B.

### Sample Collection and Processing

Whole blood and serum were collected and processed. Nasal brushings were placed in RNA Later and stored at –80°C prior to extraction for further analysis. Nasal SAM strips were frozen initially at –80°C and then were thawed on ice for processing. The absorbent strip was removed from its handle and placed into a micro centrifuge tube. Elution buffer (300 µl, 1% BSA in PBS) was added on top of the SAM strip. The tube containing the strip with buffer was vortexed for 30 seconds. Using sterile forceps, the strip was removed, a spin filter mini-column (catalog no. 9301; Costar) was inserted in the same tube, and the strip was replaced inside column. Samples were centrifuged at 16,000 x g for 20 min at 4°C. After removing samples from the centrifuge, the strip and spin column were removed. The eluate was aliquoted and stored at – 80°C.

### Immunologic Assays

Standard methods were used to measure serum HAI and NAI antibody titers against the challenge virus (42–44). ELISA for IgA and IgG antibodies against HA, NA, and HA stalk was performed on nasal secretions and serum, respectively, as previously described (45, 46) with minor modifications (Table S1).

### Calculation of IgA and IgG antibody titers against HA, NA, and HA stalk

Antibody titers were calculated using an extrapolation method. Four serum and SAM samples with the highest reactivity to each construct (HA, NA, and HA stalk) were preselected using the ELISA technique described. These samples were pooled in equal volumes and serially diluted to create the standard for its respective construct and sample type. This standard was used to generate a standard curve against the determined titer for each plate. The titer was determined by running an ELISA plate per antigen with the standard serially diluted multiple times, with various dilution factors. The OD_490_ of each dilution was measured using the ELISA method described, and the cutoff value was set as the mean optical density (OD) plus 3 standard deviations (47) of the wells containing only the secondary antibody. The titer was defined as the highest dilution factor to produce an OD value above the generated cutoff value. The titer of the mucosal sample standard was 37,500 for HA, 8,600 for NA, and 16,000 for HA stalk. The titer of the serum standard was 409,600 for HA, 12,800 for NA, and 218,000 for HA stalk. Serial dilutions of the standard were added to each plate to generate the standard curve. The constructs’ antibody titers were measured three times independently and means of the replicates were used for further analysis.

### Statistical Analysis

Participants were divided among two groups: vaccinated and unvaccinated. Wilcoxon rank sum tests were used to compare continuous outcomes between groups and within a group between timepoints. Fisher’s exact tests were used for categorical outcomes. Univariate linear regression models were used to examine the relationship between log-transformed antibody titer values and continuous outcomes of number of symptoms, number of days of symptoms, number of days of shedding, and FLU-PRO scores for each group separately, and for all subjects combined. Univariate logistic regression models were used to examine the relationship between log-transformed antibody titer values and binary outcomes of presence of Flu, symptoms, shedding, and MMID for each group separately and for all subjects combined. Spearman’s rank correlations coefficients were calculated for specified sets of antibody titers at days –1, 7, and 56. Spearman’s rank correlations coefficients were also calculated for all antibody titers between timepoints (days –1 and 7, days –1 and 56, days 7 and 56). Participants were grouped by shedding duration into three groups: non-shedders, 1-day shedders, and long-shedders (>1 day of shedding). Kruskal-Wallis tests were used to compare pre-challenge antibody titers and post-challenge symptom-based clinical outcomes between the shedding groups. An exhaustive search to find the best model fit using AIC was done using all log-transformed antibody measures at phase 2 day –1 as predictors for each outcome measure. The model with the lowest AIC was selected and the R^2^ was calculated. Groups were modeled separately. P-values were adjusted for multiple comparisons to maintain the 0.05 level of significance as detailed in the supplementary appendix. All analyses were done in R version 3.6.3. Heatmap figures were created in Prism 9.3.1.

## Data Availability

All data produced in the present study are available upon reasonable request to the authors

## Acknowledgments

This project has been funded in in part with federal funds from the National Cancer Institute, National Institutes of Health, under Contract No. 75N91019D00024 and in part with funding from the Division of Intramural Research, NIAID. The content of this publication does not necessarily reflect the views or policies of the Department of Health and Human Services, nor does mention of trade names, commercial products, or organizations imply endorsement by the U.S. Government.

## References

1. Morens DM, Taubenberger JK, Fauci AS. 2023. Rethinking next-generation vaccines for coronaviruses, influenzaviruses, and other respiratory viruses. Cell Host Microbe 31:146–157.

2. Cox R. 2013. Correlates of protection to influenza virus, where do we go from here? Human Vaccines & Immunotherapeutics 9:405–408.

3. Memoli MJ, Shaw PA, Han A, Czajkowski L, Reed S, Athota R, Bristol T, Fargis S, Risos K, Powers JH, Davey RT, Taubenberger JK. 2016. Evaluation of Antihemagglutinin and Antineuraminidase Antibodies as Correlates of Protection in an Influenza A/H1N1 Virus Healthy Human Challenge Model. mBio 7.

4. Giurgea LT, Cervantes-Medina A, Walters KA, Scherler K, Han A, Czajkowski LM, Baus HA, Hunsberger S, Klein SL, Kash JC, Taubenberger JK, Memoli MJ. 2022. Sex Differences in Influenza: The Challenge Study Experience. J Infect Dis 225:715–722.

5. Maier HE, Nachbagauer R, Kuan G, Ng S, Lopez R, Sanchez N, Stadlbauer D, Gresh L, Schiller A, Rajabhathor A, Ojeda S, Guglia AF, Amanat F, Balmaseda A, Krammer F, Gordon A. 2020. Pre-existing Antineuraminidase Antibodies Are Associated With Shortened Duration of Influenza A(H1N1)pdm Virus Shedding and Illness in Naturally Infected Adults. Clin Infect Dis 70:2290–2297.

6. Monto A, Kendal A. 1973. Effect of neuraminidase antibody on Hong Kong influenza. The Lancet 301:623–625.

7. Monto AS, Petrie JG, Cross RT, Johnson E, Liu M, Zhong W, Levine M, Katz JM, Ohmit SE. 2015. Antibody to Influenza Virus Neuraminidase: An Independent Correlate of Protection. Journal of Infectious Diseases 212:1191–1199.

8. Shvartsman YS, Zykov MP. 1976. Secretory anti-influenza immunity. Adv Immunol 22:291–330.

9. Sheikh-Mohamed S, Isho B, Chao GYC, Zuo M, Cohen C, Lustig Y, Nahass GR, Salomon-Shulman RE, Blacker G, Fazel-Zarandi M, Rathod B, Colwill K, Jamal A, Li Z, de Launay KQ, Takaoka A, Garnham-Takaoka J, Patel A, Fahim C, Paterson A, Li AX, Haq N, Barati S, Gilbert L, Green K, Mozafarihashjin M, Samaan P, Budylowski P, Siqueira WL, Mubareka S, Ostrowski M, Rini JM, Rojas OL, Weissman IL, Tal MC, McGeer A, Regev-Yochay G, Straus S, Gingras AC, Gommerman JL. 2022. Systemic and mucosal IgA responses are variably induced in response to SARS-CoV-2 mRNA vaccination and are associated with protection against subsequent infection. Mucosal Immunol 15:799–808.

10. Havervall S, Marking U, Svensson J, Greilert-Norin N, Bacchus P, Nilsson P, Hober S, Gordon M, Blom K, Klingstrom J, Aberg M, Smed-Sorensen A, Thalin C. 2022. Anti-Spike Mucosal IgA Protection against SARS-CoV-2 Omicron Infection. N Engl J Med 387:1333–1336.

11. Zuo F, Marcotte H, Hammarstrom L, Pan-Hammarstrom Q. 2022. Mucosal IgA against SARS-CoV-2 Omicron Infection. N Engl J Med 387:e55.

12. Gould VMW, Francis JN, Anderson KJ, Georges B, Cope AV, Tregoning JS. 2017. Nasal IgA Provides Protection against Human Influenza Challenge in Volunteers with Low Serum Influenza Antibody Titre. Frontiers in Microbiology 8.

13. McMahon M, Kirkpatrick E, Stadlbauer D, Strohmeier S, Bouvier NM, Krammer F. 2019. Mucosal Immunity against Neuraminidase Prevents Influenza B Virus Transmission in Guinea Pigs. mBio 10.

14. Mao T, Israelow B, Pena-Hernandez MA, Suberi A, Zhou L, Luyten S, Reschke M, Dong H, Homer RJ, Saltzman WM, Iwasaki A. 2022. Unadjuvanted intranasal spike vaccine elicits protective mucosal immunity against sarbecoviruses. Science 378:eabo2523.

15. Focosi D, Maggi F, Casadevall A. 2022. Mucosal Vaccines, Sterilizing Immunity, and the Future of SARS-CoV-2 Virulence. Viruses 14.

16. Ainai A, Tamura S-i, Suzuki T, Ito R, Asanuma H, Tanimoto T, Gomi Y, Manabe S, Ishikawa T, Okuno Y, Odagiri T, Tashiro M, Sata T, Kurata T, Hasegawa H. 2011. Characterization of neutralizing antibodies in adults after intranasal vaccination with an inactivated influenza vaccine. Journal of Medical Virology 84:336–344.

17. Suzuki T, Kawaguchi A, Ainai A, Tamura S-i, Ito R, Multihartina P, Setiawaty V, Pangesti KNA, Odagiri T, Tashiro M, Hasegawa H. 2015. Relationship of the quaternary structure of human secretory IgA to neutralization of influenza virus. Proceedings of the National Academy of Sciences 112:7809-7814.

18. Treanor J, Nolan C, O’Brien D, Burt D, Lowell G, Linden J, Fries L. 2006. Intranasal administration of a proteosome-influenza vaccine is well-tolerated and induces serum and nasal secretion influenza antibodies in healthy human subjects. Vaccine 24:254–262.

19. Ashkenazi S, Vertruyen A, Aristegui J, Esposito S, McKeith DD, Klemola T, Biolek J, Kuhr J, Bujnowski T, Desgrandchamps D, Cheng SM, Skinner J, Gruber WC, Forrest BD, Group C-TS. 2006. Superior relative efficacy of live attenuated influenza vaccine compared with inactivated influenza vaccine in young children with recurrent respiratory tract infections. Pediatr Infect Dis J 25:870–9.

20. Chung JR, Flannery B, Thompson MG, Gaglani M, Jackson ML, Monto AS, Nowalk MP, Talbot HK, Treanor JJ, Belongia EA, Murthy K, Jackson LA, Petrie JG, Zimmerman RK, Griffin MR, McLean HQ, Fry AM. 2016. Seasonal Effectiveness of Live Attenuated and Inactivated Influenza Vaccine. Pediatrics 137:e20153279.

21. Han A, Poon JL, Powers JH, 3rd, Leidy NK, Yu R, Memoli MJ. 2018. Using the Influenza Patient-reported Outcome (FLU-PRO) diary to evaluate symptoms of influenza viral infection in a healthy human challenge model. BMC Infect Dis 18:353.

22. Wang TT, Sewatanon J, Memoli MJ, Wrammert J, Bournazos S, Bhaumik SK, Pinsky BA, Chokephaibulkit K, Onlamoon N, Pattanapanyasat K, Taubenberger JK, Ahmed R, Ravetch JV. 2017. IgG antibodies to dengue enhanced for FcgammaRIIIA binding determine disease severity. Science 355:395–398.

23. 2022. Centers for Disease Control and Prevention. Past Seasons Vaccine Effectiveness Estimates. https://www.cdc.gov/flu/vaccines-work/effectiveness-studies.htm. Accessed June 1 2023.

24. Rondy M, El Omeiri N, Thompson MG, Leveque A, Moren A, Sullivan SG. 2017. Effectiveness of influenza vaccines in preventing severe influenza illness among adults: A systematic review and meta-analysis of test-negative design case-control studies. J Infect 75:381–394.

25. Atmar RL, Keitel WA, Cate TR, Munoz FM, Ruben F, Couch RB. 2007. A dose–response evaluation of inactivated influenza vaccine given intranasally and intramuscularly to healthy young adults. Vaccine 25:5367–5373.

26. Fulk RV, Fedson DS, Huber MA, Fitzpatrick JR, Howar BF, Kasel JA. 1969. Antibody responses in children and elderly persons following local or parenteral administration of an inactivated influenza virus vaccine, A2-Hong Kong-68 variant. J Immunol 102:1102–5.

27. Greenbaum E, Engelhard D, Levy R, Schlezinger M, Morag A, Zakay-Rones Z. 2004. Mucosal (SIgA) and serum (IgG) immunologic responses in young adults following intranasal administration of one or two doses of inactivated, trivalent anti-influenza vaccine. Vaccine 22:2566–77.

28. Kasel JA, Hume EB, Fulk RV, Togo Y, Huber M, Hornick RB. 1969. Antibody responses in nasal secretions and serum of elderly persons following local or parenteral administration of inactivated influenza virus vaccine. J Immunol 102:555–62.

29. Keitel WA, Cate TR, Nino D, Huggins LL, Six HR, Quarles JM, Couch RB. 2001. Immunization against influenza: comparison of various topical and parenteral regimens containing inactivated and/or live attenuated vaccines in healthy adults. J Infect Dis 183:329–332.

30. Muhamed G, Greenbaum E, Zakay-Rones Z. 2006. Neuraminidase antibody response to inactivated influenza virus vaccine following intranasal and intramuscular vaccination. Isr Med Assoc J 8:155–8.

31. Muszkat M, Greenbaum E, Ben-Yehuda A, Oster M, Yeu’l E, Heimann S, Levy R, Friedman G, Zakay-Rones Z. 2003. Local and systemic immune response in nursing-home elderly following intranasal or intramuscular immunization with inactivated influenza vaccine. Vaccine 21:1180–6.

32. Waldman RH, Kasel JA, Fulk RV, Togo Y, Hornick RB, Heiner GG, Dawkins AT, Mann JJ. 1968. Influenza Antibody in Human Respiratory Secretions after Subcutaneous or Respiratory Immunization with Inactivated Virus. Nature 218:594–595.

33. Zahradnik JM, Kasel JA, Martin RR, Six HR, Cate TR. 1983. Immune responses in serum and respiratory secretions following vaccination with a live cold-recombinant (CR35) and inactivated A/USSR/77 (H1N1) influenza virus vaccine. J Med Virol 11:277–85.

34. Giurgea LT, Morens DM, Taubenberger JK, Memoli MJ. 2020. Influenza Neuraminidase: A Neglected Protein and Its Potential for a Better Influenza Vaccine. Vaccines (Basel) 8.

35. Zost SJ, Wu NC, Hensley SE, Wilson IA. 2019. Immunodominance and Antigenic Variation of Influenza Virus Hemagglutinin: Implications for Design of Universal Vaccine Immunogens. J Infect Dis 219:S38-S45.

36. Tan HX, Jegaskanda S, Juno JA, Esterbauer R, Wong J, Kelly HG, Liu Y, Tilmanis D, Hurt AC, Yewdell JW, Kent SJ, Wheatley AK. 2019. Subdominance and poor intrinsic immunogenicity limit humoral immunity targeting influenza HA stem. J Clin Invest 129:850–862.

37. Cohen JI, Dropulic L, Wang K, Gangler K, Morgan K, Liepshutz K, Krogmann T, Ali MA, Qin J, Wang J, Vogel JS, Lei Y, Suzuki-Williams LP, Spalding C, Palmore TN, Burbelo PD. 2023. Comparison of Levels of Nasal, Salivary, and Plasma Antibody to Severe Acute Respiratory Syndrome Coronavirus 2 During Natural Infection and After Vaccination. Clin Infect Dis 76:1391–1399.

38. Wright PF, Prevost-Reilly AC, Natarajan H, Brickley EB, Connor RI, Wieland-Alter WF, Miele AS, Weiner JA, Nerenz RD, Ackerman ME. 2022. Longitudinal Systemic and Mucosal Immune Responses to SARS-CoV-2 Infection. J Infect Dis 226:1204–1214.

39. Johansson BE, Moran TM, Kilbourne ED. 1987. Antigen-presenting B cells and helper T cells cooperatively mediate intravirionic antigenic competition between influenza A virus surface glycoproteins. Proc Natl Acad Sci U S A 84:6869–73.

40. Clements ML, Betts RF, Tierney EL, Murphy BR. 1986. Serum and nasal wash antibodies associated with resistance to experimental challenge with influenza A wild-type virus. J Clin Microbiol 24:157–60.

41. Memoli MJ, Czajkowski L, Reed S, Athota R, Bristol T, Proudfoot K, Fargis S, Stein M, Dunfee RL, Shaw PA, Davey RT, Taubenberger JK. 2015. Validation of the wild-type influenza A human challenge model H1N1pdMIST: an A(H1N1)pdm09 dose-finding investigational new drug study. Clin Infect Dis 60:693–702.

42. Cottey R, Rowe CA, Bender BS. 2001. Influenza virus. Curr Protoc Immunol Chapter 19:Unit 19.11.

43. Potter CW, Oxford JS. 1979. Determinants of immunity to influenza infection in man. Br Med Bull 35:69–75.

44. Wan H, Gao J, Xu K, Chen H, Couzens LK, Rivers KH, Easterbrook JD, Yang K, Zhong L, Rajabi M, Ye J, Sultana I, Wan XF, Liu X, Perez DR, Taubenberger JK, Eichelberger MC. 2013. Molecular basis for broad neuraminidase immunity: conserved epitopes in seasonal and pandemic H1N1 as well as H5N1 influenza viruses. J Virol 87:9290–300.

45. Park JK, Xiao Y, Ramuta MD, Rosas LA, Fong S, Matthews AM, Freeman AD, Gouzoulis MA, Batchenkova NA, Yang X, Scherler K, Qi L, Reed S, Athota R, Czajkowski L, Han A, Morens DM, Walters KA, Memoli MJ, Kash JC, Taubenberger JK. 2020. Pre-existing immunity to influenza virus hemagglutinin stalk might drive selection for antibody-escape mutant viruses in a human challenge model. Nat Med 26:1240–1246.

46. Park JK, Han A, Czajkowski L, Reed S, Athota R, Bristol T, Rosas LA, Cervantes-Medina A, Taubenberger JK, Memoli MJ. 2018. Evaluation of Preexisting Anti-Hemagglutinin Stalk Antibody as a Correlate of Protection in a Healthy Volunteer Challenge with Influenza A/H1N1pdm Virus. mBio 9.

47. Layne SP, Beugelsdijk TJ, Patel CK, Taubenberger JK, Cox NJ, Gust ID, Hay AJ, Tashiro M, Lavanchy D. 2001. A global lab against influenza. Science 293:1729.

